# Impact of a Phased Transition Model on Advanced HIV Disease Outcomes: A Pre- and Post-Implementation Evaluation Study in Malawi

**DOI:** 10.64898/2026.04.13.26350558

**Authors:** Thulani Maphosa, Rhoderick Machekano, Lise Denoeud-Ndam, Lucky Makonokaya, Lloyd Chilikutali, Louiser Upile Kalitera, Eddington Matiya, Allan Mayi, Reuben Musarandega, Bilal Wilson Matola, Aida Yemane Berhan, Allan Ahimbisibwe, Appolinaire Tiam

## Abstract

**Background:** To promote sustainability and strengthen national ownership of Advanced HIV Disease (AHD) services, a transition was implemented across 22 health facilities in Central Malawi. This transition involved shifting responsibility for key AHD program elements, including clinical service delivery, diagnostics, provider mentorship, and reporting systems, from implementing partner-led implementation to full Ministry of Health (MoH) leadership. This evaluation assessed the impact of this transition on diagnostic coverage, TB preventive therapy (TPT) uptake, and 12-month survival outcomes.

**Methods:** A retrospective cohort study was conducted involving all children and adults enrolled in AHD care during the pre-MoH transition (January 2020–December 2021) and post-MoH transition (January 2023–December 2024) periods. Eligibility followed national AHD criteria: CD4 count <200 cells/mm^3^, WHO stage 3 or 4 illness, or age <5 years. AHD clients’ data were abstracted from clinical records and linked across routine facility registers to assess diagnostic and treatment indicators. Kaplan–Meier survival curves, Cox proportional hazards, and Fine and Gray competing risk models were used to evaluate 6 and 12-month mortality and retention as primary outcomes.

**Results:** A total of 1,044 AHD clients were included (553 pre-transition; 491 post-transition) in the evaluation. Median age increased post-transition (35.9 to 38.5 years, p<0.001). CD4 testing declined (80.7% to 46.0%, p<0.001) testing uptake, while WHO staging and TB diagnostic coverage improved. TB diagnoses decreased (44.5% to 31.2%, p=0.002). TPT uptake dropped from 46.4% to 31.6% (p<0.001). Twelve-month mortality significantly declined from 9.4% to 5.5% (adjusted hazard ratio [aHR]=0.59, 95% CI: 0.37–0.94, p=0.026). Retention in care remained stable (HR=0.86, 95% CI: 0.62– 1.20, p=0.384).

**Conclusions:** Transitioning AHD services to MoH leadership sustained key program outcomes and significantly reduced mortality. Continued mentorship and government ownership were key drivers of success. However, declines in CD4 testing and TPT coverage highlight the need for strengthened diagnostics and preventive care integration. These findings support scaling nationally-led AHD models in high-burden HIV settings.

## Background

Despite substantial progress in global HIV care and treatment, Advanced HIV Disease (AHD) remains a persistent contributor to AIDS-related morbidity and mortality, particularly in sub-Saharan Africa(1-3). Nearly one in three people starting antiretroviral therapy (ART) in the region presents with advanced disease, defined by the WHO as having a CD4 count <200 cells/mm^3^ or WHO stage 3 or 4 clinical disease or age <5 years. In Malawi, a country with a high HIV burden and longstanding ART programs, approximately 50% of adult ART clients with AHD are not diagnosed or managed appropriately, especially in decentralized settings(3-5). This gap results in late detection of opportunistic infections such as tuberculosis (TB), cryptococcal meningitis, and severe bacterial infections, which account for the majority of preventable HIV-related deaths(1, 3, 6-8).

In response, the World Health Organization developed the AHD care package, recommending a standardized approach that includes rapid CD4 testing, TB-lipoarabinomannan (LAM) and cryptococcal antigen (CrAg) screening, and expedited ART initiation(2). However, most AHD program implementation has been partner-led and concentrated at central hospitals, creating a sustainability gap in health system ownership, capacity, and integration. In Malawi, between 2020 and 2021, Elizabeth Glaser Pediatric AIDS Foundation (EGPAF), a non-governmental organization (NGO), introduced a hub-and-spoke model at 22 health hospitals, providing diagnostics, training, mentorship, and treatment support for AHD. While this initiative yielded improvements in detection and outcomes, questions remained around how to sustain these interventions as donor support wanes and national health systems are expected to assume full ownership(9-11).

To address this, a structured transition model was developed and implemented beginning in 2022 to transfer full responsibility for AHD service delivery to the Malawi Ministry of Health (MoH). The transition encompassed key domains of service delivery, including commodity management, health worker training, mentorship, data integration, and supervision, shifting leadership from EGPAF to district health offices and national HIV program units. This transition was critical to institutionalizing AHD care within existing MoH systems, ensuring continuity and sustainability of lifesaving interventions(11, 12).

We conducted an evaluation to document the feasibility, effectiveness, and outcomes of transitioning a complex HIV service delivery model from partner-led to nationally owned. Most evaluations focus on clinical outcomes or short-term donor performance metrics. Few have rigorously examined what happens to program coverage, quality, and patient outcomes after such a transition, particularly for high-risk populations like those with AHD. By understanding these dynamics, national HIV programs and development partners can design more resilient systems and optimize resource allocation to sustain progress toward the UNAIDS 95-95-95 goals and reduce AIDS-related mortality(2).

To assess the implementation and outcomes of the AHD program transition in Malawi, we evaluated (1) whether health service coverage, clinical outcomes, and patient retention among clients with AHD changed significantly between the pre-transition (2020–2021) and post-transition (2022–2023) periods, and (2) the extent to which the MoH successfully assumed responsibility for program delivery, supervision, and data integration across transition domains.

## Methods

### Study Design and Setting

We conducted a non-randomised, cluster-based pre-post implementation study to evaluate the outcomes of transitioning an optimized AHD care model from a partner-led approach to full MoH ownership in Malawi. The intervention was implemented in three districts, Ntcheu, Dedza, and Mchinji, in the Central Region of Malawi, selected due to their high HIV burden and early adoption of the AHD care package. The evaluation study was implemented across 22 purposively selected health facilities, comprising 10 hub hospitals and 12 satellite spoke sites. The intervention was implemented in three temporal phases aligned with the progressive transition of responsibilities from partner-led to government-led management. Table 1 shows that during the pre-transition phase (January 2020 to December 2021), the program was led by the EGPAF, which provided full implementation support. This was followed by the transition phase (January to December 2022), during which EGPAF and the MoH jointly coordinated activities to facilitate capacity-building and systems integration. Finally, in the post-transition phase (January 2023 to December 2024), full program leadership and operations were assumed by the MoH. Client outcomes across all phases were assessed using a retrospective cohort design with follow-up extending up to 12 months after enrolment into AHD care.

**Table 1:** Intervention Description for AHD Program Transition Matrix.

| Domain | Pre-Transition (2020–2021) | Transition Phase (2022) | Post-Transition (2023–2024) |
| --- | --- | --- | --- |
| **Program Leadership** | Externally driven implementation led by EGPAF | Co-governance established between EGPAF and MOH | Full institutional ownership and leadership by MOH |
| **AHD Diagnostic Commodities** | Parallel procurement and distribution managed by EGPAF | Strengthened stock visibility through integration with MOH LMIS | Fully integrated procurement within national supply chain systems |
| **Health Workforce Capacity** | Facility-based training led by EGPAF | Embedded mentorship co-delivered with MOH teams | Institutionalized MOH-led CME and supportive supervision systems |
| **Service Delivery Model** | Centralized delivery at 10 hub facilities | Decentralization to spoke facilities with partner support | District-wide scale-up across 22 facilities under MOH |
| **Monitoring & Evaluation Systems** | Parallel AHD registers and reporting systems | Progressive alignment with national reporting frameworks | Full integration into national DHIS2 architecture |
| **Guidelines & SOPs** | Partner-developed job aids and protocols | Joint revision and contextualization with MOH input | Nationally endorsed and standardized AHD SOPs |
| **Mentorship & Technical Assistance** | Intensive partner-led technical assistance | Transition to joint mentorship through MOH TWGs | MOH-led mentorship systems with minimal external support |
| **Community Linkages** | Limited, facility-based patient identification | Pilot community referral pathways initiated | Integration with CHWs and Expert Clients within national programs |
| **Evidence Generation & Use** | Partner-driven operational research | Structured dissemination to MOH TWGs and stakeholders | Routine MOH-led data review and evidence-informed decision-making |
| **Quality Improvement Systems** | Partner-led QI data collection and review | Introduction of joint QI coaching and learning platforms | Institutionalized MOH-led DQA and continuous QI systems |

**Notes:** EGPAF, Elizabeth Glaser Pediatric AIDS Foundation; **MoH**, Ministry of Health; **LMIS**, Laboratory Management Information System; **CME**, Continuing Medical Education; **SOP**, Standard Operating Procedure; **TWG**, Technical Working Group; **CHW**, Community Health Worker; **DQA**, Data Quality Assessment, **QI**, Quality Improvement.

### Study Population and Eligibility Criteria

The study population comprised all children and adults enrolled in AHD care at the 22 study sites during the pre- and post-transition phases (excluding year 2022 when transition was ongoing). Eligibility for inclusion was based on Malawi’s national AHD criteria, which included people living with HIV with a CD4 count of less than 200 cells/mm^3^, a WHO clinical stage 3 or 4 illness, or being under 5 years of age, regardless of clinical or immunologic status. Clients were excluded if they lacked documented AHD-defining criteria or had incomplete clinical records that could not be verified. In total, 1,044 clients were included in the final analysis, 553 during the pre-transition phase and 491 during the post-transition phase.

### Intervention Description

The intervention involved the structured transition of responsibilities across nine key domains: program leadership, commodity procurement, human resource capacity, service site coverage, monitoring and evaluation, guidelines and SOPs, mentorship and technical assistance, community linkages, and quality improvement.

The pre-transition phase was characterized by intensive EGPAF support, including partner-led procurement, training, and parallel monitoring systems. During the transition phase, joint planning and capacity-building with the MoH were implemented. In the post-transition phase, all program components were fully managed by the MoH, with integration into national systems such as DHIS2 and health facility-led mentorship. **Table 1**.

### Outcomes and Follow-up

The primary outcomes for this study were all-cause mortality and retention up to 12 months following enrolment into AHD care. Secondary outcomes included retention in care at 3, 6, and 12 months; uptake of TB preventive therapy (TPT) among eligible clients; diagnostic coverage encompassing CD4 testing, CrAg screening, and GeneXpert testing for TB; as well as the frequency of opportunistic infections at the time of presentation. All clients enrolled in the cohort were followed for a period of up to 12 months from the date of AHD enrolment to assess these outcomes.

### Sample Size and Power

Assuming a baseline 6-month mortality of 12%, we calculated that a minimum of 440 clients per study phase would be required to detect a 30% relative reduction in mortality (α = 0.05, power = 80%). The achieved sample (n = 1,044) provided adequate statistical power for subgroup and survival analyses.

### Data Collection and Management

Data were retrospectively extracted from multiple sources, including the AHD register, ART patient files, laboratory logs, and the national EMR system. Data from the pre-intervention collection started on 2 November 2021, while for the post-intervention on 16 February 2024, ART numbers served as unique identifiers to triangulate and de-duplicate records. Data abstraction was conducted using Open Data Kit (ODK) tablets, and datasets were exported into Stata 17.0 (StataCorp, College Station, TX) for analysis.

### Statistical Analysis

Descriptive statistics were used to summarize demographic and clinical characteristics. Pre-post differences were assessed using Chi-square or Fisher’s exact tests for categorical variables and Wilcoxon rank-sum tests for non-parametric continuous variables.

Kaplan–Meier survival curves and the log-rank test were used to compare time-to-death and retention outcomes. Cox proportional hazards models and Fine-Gray sub-distribution hazard models were applied to assess the effect of the intervention while adjusting for potential confounders and accounting for competing risks such as treatment stoppage and transfer out. Results are presented as hazard ratios (HRs) and sub-distribution hazard ratios (sHRs) with 95% confidence intervals. A p-value <0.05 was considered statistically significant.

## Results

Among the 1,044 clients enrolled in AHD care, baseline characteristics varied significantly between the pre- and post-transition periods (**Table 2**). There was a comparable sex distribution across phases (p=0.40), with females constituting slightly more than half of the population. However, age distributions differed significantly (p=0.007), with a greater proportion of older clients (≥50 years) enrolled post-transition. The mean age was also significantly higher post-transition (35.9 vs. 38.5 years, p<0.001).

**Table 2.**
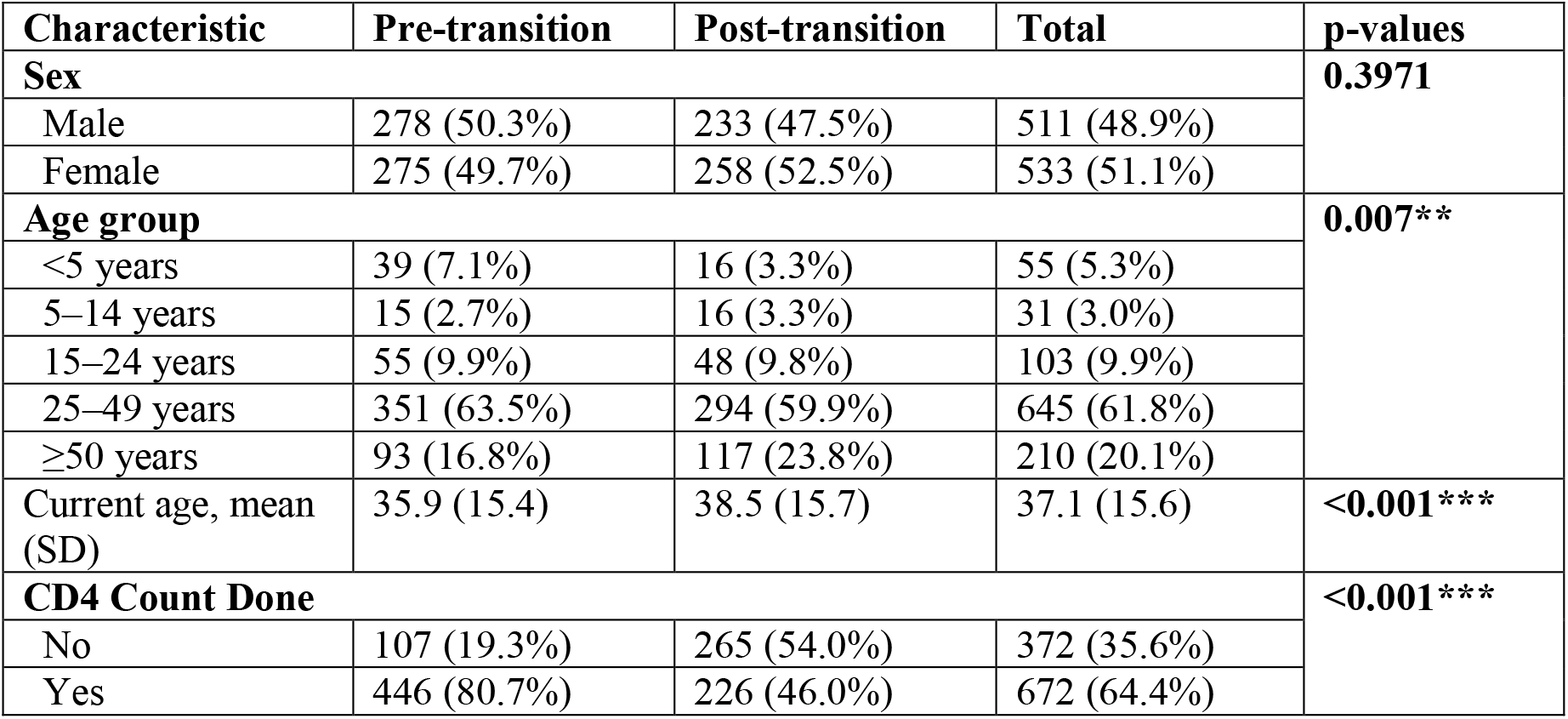
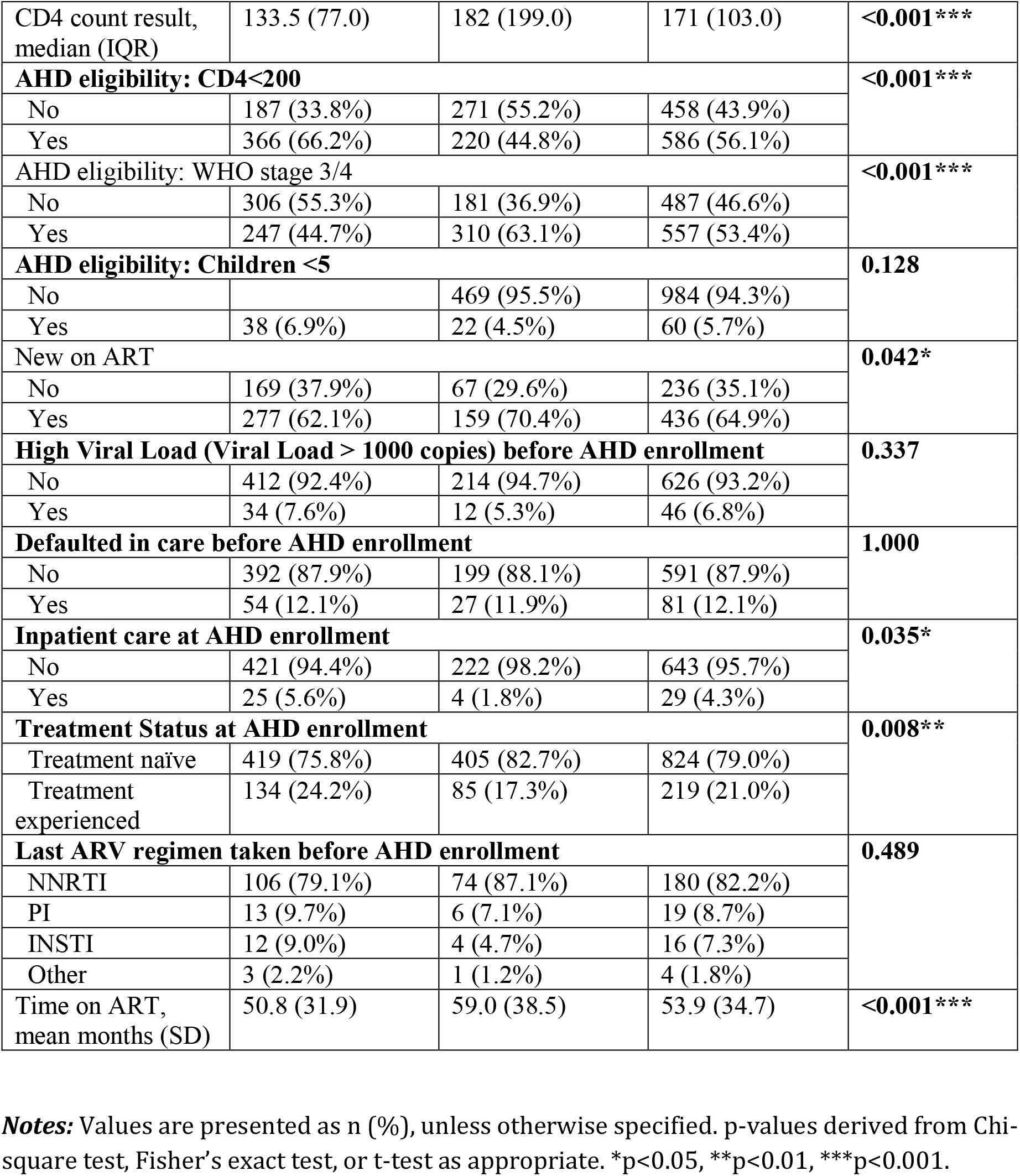
Baseline Characteristics of Clients by Phase of AHD Program Implementation.

CD4 testing coverage declined pre-to post-transition (80.7% vs. 46.6%, respectively, p<0.001), although the median CD4 count was higher in the post-than pre-transition group (134 vs. 182, respectively, p<0.001) (**Table 2**). A higher proportion of clients met AHD eligibility based on CD4<200 during the pre-than post-transition period (66.2% vs. 44.8%, respectively, p<0.001), while more were classified as AHD based on WHO stage 3/4 in the post- than pre-transition phase (44.7% vs. 63.1%, respectively, p<0.001). The proportion of new ART initiates was higher post-than pre-transition phase (62.1% vs 70.4%, p=0.04), and fewer clients at post-transition were enrolled as inpatients (5.6% vs. 1.8%, p=0.0348). Treatment-naïve clients were more in post- than pre-transition phase (75.8% vs. 82.7%, respectively, p=0.008), and the mean time on ART was significantly longer in post-transition (50.8 vs. 59.0 months, p<0.001). No significant differences were observed in viral load suppression, ART defaulting before AHD enrollment, or prior ART regimens.

**Table 3** highlights key differences in the clinical presentation of clients across the AHD program phases. TB remained the most frequently diagnosed opportunistic infection but showed a significant decline from 44.5% in the pre-transition period to 31.2% post-transition (p = 0.002). Similarly, significant reductions were observed in cases of candidiasis (3.4% vs. 0.0%, p = 0.005) and severe weight loss (12.5% vs. 5.0%, p = 0.003). In contrast, bacterial infections increased significantly in the post-compared to the pre-transition phase (6.8% to 12.5%, respectively, p = 0.04). Other conditions, such as cryptococcal meningitis, Kaposi sarcoma, and severe pneumonia, were less common and did not differ significantly between phases.

**Table 3.**
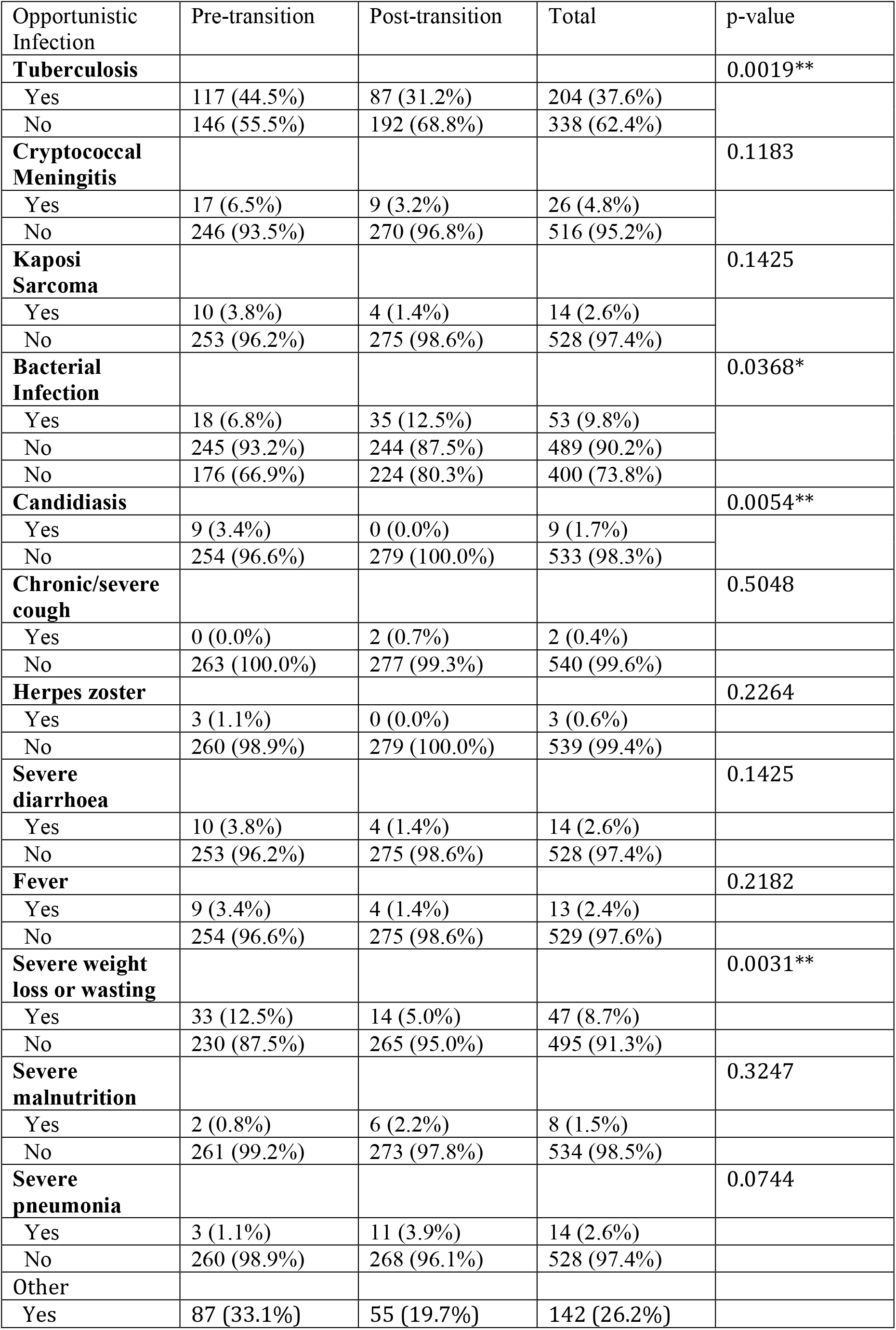
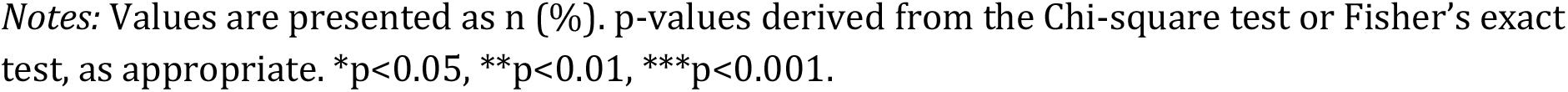
Opportunistic Infections and Related Signs and Symptoms by phase of AHD Program Implementation.

| Opportunistic Infection | Pre-transition | Post-transition | Total | p-value |
| --- | --- | --- | --- | --- |
| **Tuberculosis** |  |  |  | 0.0019** |
| Yes | 117 (44.5%) | 87 (31.2%) | 204 (37.6%) |  |
| No | 146 (55.5%) | 192 (68.8%) | 338 (62.4%) |  |
| **Cryptococcal Meningitis** |  |  |  | 0.1183 |
| Yes | 17 (6.5%) | 9 (3.2%) | 26 (4.8%) |  |
| No | 246 (93.5%) | 270 (96.8%) | 516 (95.2%) |  |
| **Kaposi Sarcoma** |  |  |  | 0.1425 |
| Yes | 10 (3.8%) | 4 (1.4%) | 14 (2.6%) |  |
| No | 253 (96.2%) | 275 (98.6%) | 528 (97.4%) |  |
| **Bacterial Infection** |  |  |  | 0.0368* |
| Yes | 18 (6.8%) | 35 (12.5%) | 53 (9.8%) |  |
| No | 245 (93.2%) | 244 (87.5%) | 489 (90.2%) |  |
| No | 176 (66.9%) | 224 (80.3%) | 400 (73.8%) |  |
| **Candidiasis** |  |  |  | 0.0054** |
| Yes | 9 (3.4%) | 0 (0.0%) | 9 (1.7%) |  |
| No | 254 (96.6%) | 279 (100.0%) | 533 (98.3%) |  |
| **Chronic/severe cough** |  |  |  | 0.5048 |
| Yes | 0 (0.0%) | 2 (0.7%) | 2 (0.4%) |  |
| No | 263 (100.0%) | 277 (99.3%) | 540 (99.6%) |  |
| **Herpes zoster** |  |  |  | 0.2264 |
| Yes | 3 (1.1%) | 0 (0.0%) | 3 (0.6%) |  |
| No | 260 (98.9%) | 279 (100.0%) | 539 (99.4%) |  |
| **Severe diarrhoea** |  |  |  | 0.1425 |
| Yes | 10 (3.8%) | 4 (1.4%) | 14 (2.6%) |  |
| No | 253 (96.2%) | 275 (98.6%) | 528 (97.4%) |  |
| **Fever** |  |  |  | 0.2182 |
| Yes | 9 (3.4%) | 4 (1.4%) | 13 (2.4%) |  |
| No | 254 (96.6%) | 275 (98.6%) | 529 (97.6%) |  |
| **Severe weight loss or wasting** |  |  |  | 0.0031** |
| Yes | 33 (12.5%) | 14 (5.0%) | 47 (8.7%) |  |
| No | 230 (87.5%) | 265 (95.0%) | 495 (91.3%) |  |
| **Severe malnutrition** |  |  |  | 0.3247 |
| Yes | 2 (0.8%) | 6 (2.2%) | 8 (1.5%) |  |
| No | 261 (99.2%) | 273 (97.8%) | 534 (98.5%) |  |
| **Severe pneumonia** |  |  |  | 0.0744 |
| Yes | 3 (1.1%) | 11 (3.9%) | 14 (2.6%) |  |
| No | 260 (98.9%) | 268 (96.1%) | 528 (97.4%) |  |
| **Other** |  |  |  |  |
| Yes | 87 (33.1%) | 55 (19.7%) | 142 (26.2%) |  |

Notes: Values are presented as n (%). p-values derived from the Chi-square test or Fisher's exact test, as appropriate. \*p<0.05, \*\*p<0.01, \*\*\*p<0.001.

Of the 1,044 clients enrolled, 80.7% were eligible for TPT. Uptake was significantly higher in the pre-transition phase (46.4%) compared to the post-transition phase (31.6%) (p < 0.001), indicating reduced coverage following the transition (**Table 4)**.

**Table 4.**
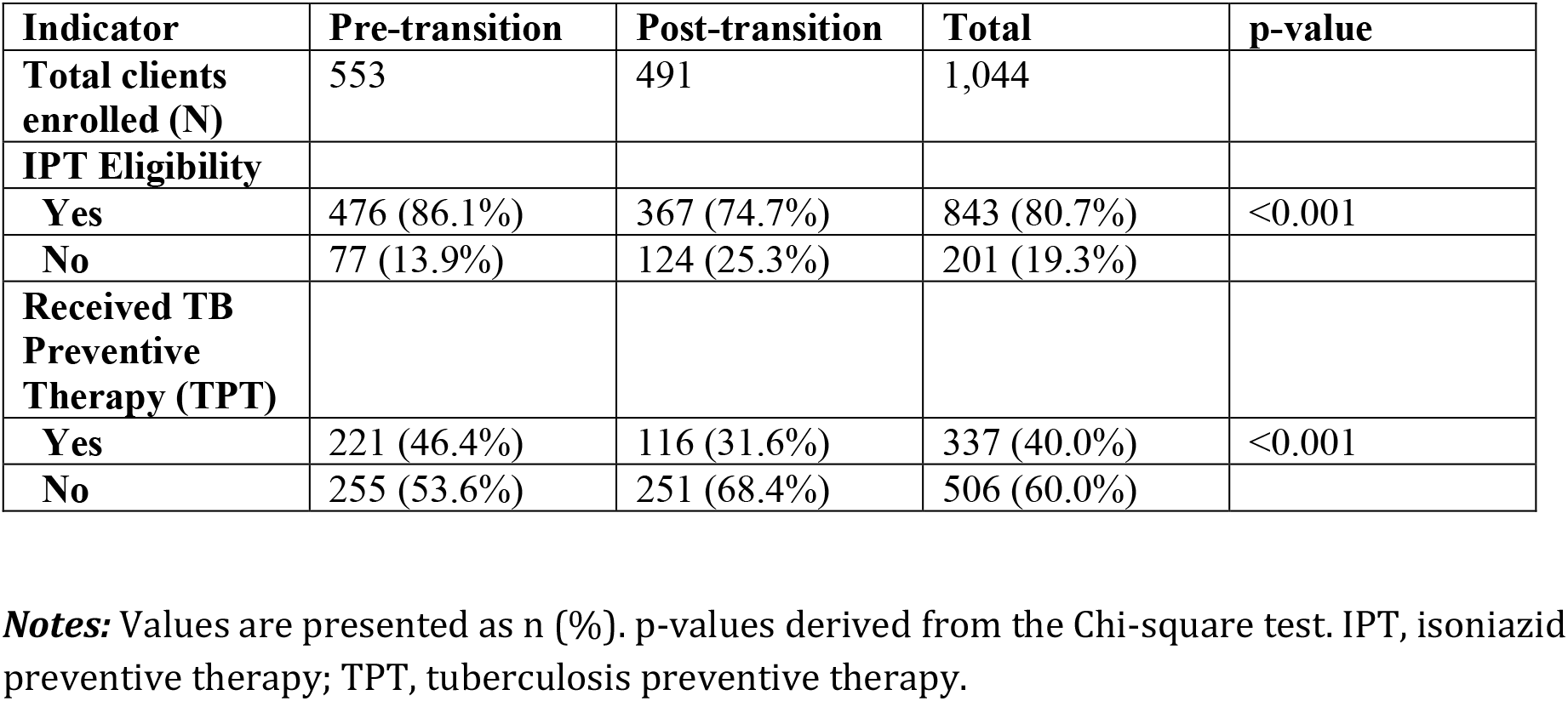
TB Preventive Therapy Among Eligible Patients by Phase of AHD Program Implementation.

| Indicator | Pre-transition | Post-transition | Total | p-value |
| --- | --- | --- | --- | --- |
| Total clients enrolled (N) | 553 | 491 | 1,044 |  |
| IPT Eligibility |  |  |  |  |
| Yes | 476 (86.1%) | 367 (74.7%) | 843 (80.7%) | <0.001 |
| No | 77 (13.9%) | 124 (25.3%) | 201 (19.3%) |  |
| Received TB Preventive Therapy (TPT) |  |  |  |  |
| Yes | 221 (46.4%) | 116 (31.6%) | 337 (40.0%) | <0.001 |
| No | 255 (53.6%) | 251 (68.4%) | 506 (60.0%) |  |

Notes: Values are presented as n (%). p-values derived from the Chi-square test. IPT, isoniazid preventive therapy; TPT, tuberculosis preventive therapy.

Treatment outcomes were comparable across phases, with 70.2% of clients alive on treatment in the pre-transition phase and 71.0% in the post-transition phase **(Table 5A)**. However, mortality was lower post-transition (9.4% vs. 5.5%), and survival analysis showed significantly reduced mortality risk in the post-transition phase at 3, 6, and 12 months **(Table 5B)** and **(Figure 1)**, supported by Cox regression (HR = 0.59, 95% CI: 0.37–0.94, *p* = 0.026) and Fine and Gray models (HR = 0.58, 95% CI: 0.36–0.93, *p* = 0.024) **(Table 5C)**. While out-of-care risk estimates slightly favored the post-transition phase at each time point **(Table 5D)**, the difference was not statistically significant (HR = 0.86, 95% CI: 0.62– 1.20, *p* = 0.384) **(Table 5E)** and **(Figure 2)**.

**Table 5:** Treatment Outcomes and Survival by Pre- and Post-Transition Period.

**A. Treatment Outcomes by Phase of AHD Program Implementation**
| Outcome | Pre-transition (n=553) | Post-transition (n=491) | p-value |
| --- | --- | --- | --- |
| Alive on treatment | 388 (70.2%) | 348 (71.0%) | **0.063** |
| Died | 52 (9.4%) | 27 (5.5%) |  |
| Transfer Out | 83 (15.0%) | 81 (16.5%) |  |
| Stopped Treatment | 2 (0.4%) | 0 (0.0%) |  |
| Defaulted | 28 (5.1%) | 34 (6.9%) |  |

B. Estimated Mortality Risk at 3, 6, and 12 Months
| Phase | Time (months) | At risk | Failures | Failure (%) | 95% CI |
| --- | --- | --- | --- | --- | --- |
| Pre-transition | 3 | 442 | 9 | 6.3% | 4.5% - 8.9% |
|  | 6 | 411 | 12 | 8.3% | 6.1% - 11.1% |
|  | 12 | 94 | 0 | 12.1% | 9.2% - 15.8% |
| Post-transition | 3 | 397 | 3 | 4.1% | 2.6% - 6.4% |
|  | 6 | 373 | 4 | 4.8% | 3.2% - 7.3% |
|  | 12 | 67 | 2 | 6.0% | 4.1% - 8.8% |

C. Comparison of Mortality Risk: Pre- vs Post-transition
| Model | Hazard Ratio | 95% CI | p-value |
| --- | --- | --- | --- |
| Cox regression | 0.59 | [0.37 - 0.94] | 0.026 |
| Fine & Gray regression | 0.58 | [0.36 - 0.93] | 0.024 |

D. Estimated Out-of-Care Risk at 3, 6, and 12 Months
| Phase | Time (months) | At risk | Failures | Failure (%) | 95% CI |
| --- | --- | --- | --- | --- | --- |
| Pre-transition | 3 | 442 | 16 | 9.0% | 6.8% - 11.9% |
|  | 6 | 411 | 18 | 12.4% | 9.8 - 15.6% |
|  | 12 | 94 | 0 | 17.6% | 14.2% - 21.6% |
| Post-transition | 3 | 397 | 10 | 8.6% | 6.3% - 11.5% |
|  | 6 | 373 | 9 | 10.9% | 8.3% - 14.2% |
|  | 12 | 67 | 3 | 13.3% | 10.4% - 16.9% |

E. Comparison of Retention Risk: Pre- vs Post-transition
| Model | Hazard Ratio | 95% CI | p-value |
| --- | --- | --- | --- |
| Cox regression | 0.86 | [0.62 - 1.20] | 0.384 |

**Notes:** Values are presented as n (%), unless otherwise indicated. CI, confidence interval. p-values derived from Chi-square test, Cox proportional hazards regression, or Fine & Gray regression, as appropriate. \*p<0.05 is considered statistically significant.

## Discussion

In this retrospective cohort study evaluating the phased transition of an AHD care model from EGPAF-led delivery to full MoH stewardship across three districts in Central Malawi, we observed a marked shift towards earlier enrolment and improved outcomes. Patients entering care in the post-transition phase presented with fewer severe opportunistic conditions, including lower prevalence of severe weight loss and candidiasis, and experienced reduced mortality compared with the NGO-led phase(11).

These findings suggest that decentralised, government-led models can not only sustain but potentially enhance program performance when supported by effective integration into routine health systems(13). They also align with global evidence underscoring that earlier diagnosis, streamlined service delivery, and program ownership by the MoH are critical determinants of survival for people with AHD(14).

The observed decline in baseline CD4 testing coverage post-transition signals a critical vulnerability in diagnostic infrastructure when donor-supported systems are integrated into routine care. However, the associated shift toward higher median CD4 counts and increased outpatient initiation suggests that case-finding improved via earlier clinical engagement, even as laboratory capacity contracted. This contrast between weakened diagnostics and more proactive care delivery is not unique; studies from low-income settings report that donor transition can erode diagnostic quality, supply chains, and service comprehensiveness, despite preserving service volume. Sustaining gains in HIV care during transition thus demands deliberate support for laboratory systems and innovative strategies, such as affordable point-of-care CD4 technologies, which maintain immunological staging capacity even in decentralized settings. Embedding such diagnostics alongside structured capacity-building and mentorship may protect early identification of AHD, preserve outpatient care benefits, and ultimately support sustainable AHD responses under government stewardship(15).

The decline in candidiasis and severe weight loss among clients enrolled post-compared to pre-transition signals earlier engagement in AHD care and a shift toward less advanced disease at presentation. These findings align with global evidence that decentralised and government-led models, when coupled with rapid ART initiation and systematic screening, can reduce the burden of severe opportunistic conditions(2, 16). However, the persistent presence of cryptococcal meningitis highlights an enduring gap in diagnostic and preventive coverage. Despite WHO recommendations for systematic CrAg screening and pre-emptive antifungal therapy, implementation remains inconsistent in routine settings, with missed opportunities for mortality prevention(2, 17). The concurrent rise in bacterial infections post-transition further reflects weaknesses in antimicrobial stewardship and limited microbiological diagnostic capacity (Kourí et al., 2022). Together, these trends underscore that while program transition has facilitated earlier and less severe disease presentation, sustaining gains requires deliberate investment in laboratory strengthening, full implementation of the AHD package, including CrAg screening and integrated infection management strategies to prevent reversals in progress.

The decrease in mortality with aHR of 0.59 and 0.58 in Cox and Fine–Gray models, respectively, demonstrates the critical value of structured capacity-building and government-led mentorship in stabilizing high-risk AHD cohorts. Notably, this mortality reduction occurred without a corresponding decline in retention, indicating that strengthened clinical outcomes were not offset by increased disengagement from care. These findings are consistent with an expanding body of evidence showing that when complex HIV interventions are transitioned into national health systems, outcomes improve through institutionalized mentoring, workforce strengthening, and integrated service delivery(9, 14, 18, 19). Moreover, embedding AHD management into government platforms aligns with implementation research that underscores sustainability as a function of local ownership, capacity transfer, and system integration(14). Together, these results affirm that a government-led AHD strategy, supported by deliberate capacity-building and robust follow-up systems, can achieve measurable survival benefits while laying the foundation for long-term program sustainability.

However, the observed decline in TPT uptake, from 46% to 32% during the transition period, is concerning, as it highlights how programmatic realignments can inadvertently disrupt the continuity of preventive care. Even though reductions in eligibility mirrored declines in diagnostic uptake, this pattern exposes the inherent fragility of TB prevention services in decentralized, transitioning settings. Implementation research underscores that maintaining TPT delivery during stewardship transitions demands robust systems for commodity forecasting, integrated monitoring, and proactive follow-up mechanisms; without these, preventive gains quickly erode(20-22). Importantly, safeguards such as task-sharing with community health workers, home-based initiation and follow-up, and integration of TPT into broader primary care platforms have demonstrated effectiveness in preserving preventive coverage in decentralized contexts. As such, national policies must prioritize embedding these systemic supports, particularly during program transitions to sustain progress in AHD interventions and protect the downstream impact on HIV-related mortality(12). From a health systems perspective, our transition model, with comprehensive matrices covering leadership, procurement, training, monitoring, SOPs, and community linkage, provides a replicable blueprint for program ownership by the governments. Unlike short-term donor-driven projects with limited institutionalization, our phased approach yielded sustained coverage of essential services post-handover, evidenced by mortality benefit supporting calls for integrating evidence generation and co-accountability during handovers.

Several limitations warrant acknowledgment. The observational design precludes causal inference. Documentation gaps and AHD-related supply pipelines, particularly CD4 usage, may have biased baseline AHD definitions. Additionally, we did not quantify cost or measure intervention fidelity, which are important given resource constraints in low-income settings. Despite this, our large real-world dataset, rigorous 12-month follow-up, and stratified survival and competing-risk analyses strengthen the credibility and policy relevance of our findings.

In conclusion, this study demonstrates that a structured and phased transition of AHD services to national stewardship can deliver sustained mortality reductions without eroding retention, underscoring the feasibility of embedding complex care models within routine health systems. Success was contingent on deliberate capacity-building, robust monitoring, and high-level stakeholder alignment, critical pillars for resilient health system integration. These findings are directly relevant to global HIV strategies seeking to re-centre program ownership within MoH, advancing both sustainability and equity in line with calls to “decolonise” global health. Future priorities must include rigorous evaluation of cost-effectiveness, fidelity to evidence-based clinical algorithms, and scalable delivery models, ensuring that the gains demonstrated here can be replicated across sub-Saharan Africa and other high-burden settings.

## Data Availability

Data Availability Statement The data underlying the results presented in this study are not publicly available due to ethical and legal restrictions, as they contain potentially identifiable patient-level information collected from routine clinical records within the Malawi Ministry of Health system. Data are available upon reasonable request from the corresponding author, and with permission from the Malawi Ministry of Health and the Elizabeth Glaser Pediatric AIDS Foundation. Requests for data access will be subject to review by the relevant ethical and regulatory bodies to ensure compliance with data protection and confidentiality requirements.

## DECLARATIONS

### Ethics approval and consent to participate

Data collection was limited to secondary data abstraction of data that were routinely collected and documented as part of the AHD standard medical care of the project, without any direct interaction between the program evaluation staff and study participants. This study and all the procedures involved in collecting and analyzing routine program data imposed minimal risk to participants; therefore, the Institutional Review Boards granted a waiver of consent.

Ethical clearance was obtained from the National Health Science Research Committee on 21/06/2720. The Advarra Institutional Review Board reviewed and approved the study on 6 October 2021 (protocol number Pro00057717). All study methods were performed in accordance with relevant guidelines and regulations.

### Consent for publication

Not applicable

### Availability of data and materials

Data are available on reasonable request. Data may be obtained from a third party and is not publicly available.

### Authors’ contributions

TM, AT, LD conceptualized and designed the study, including the development of the research questions and methodology. TM, LD, LC, BW, EM, and RC were responsible for overseeing data collection, ensuring data quality, and maintaining data integrity throughout the process. RM conducted the data analysis, applying appropriate statistical techniques and interpreting the results. TM took the lead in drafting the manuscript, including writing the initial draft and subsequent revisions. TM, AT, and LDN provided critical revisions for important intellectual content and contributed to the interpretation of the findings. TM, LC, and RC contributed to the review and editing of the manuscript, ensuring clarity and coherence. RM provided substantial input on the data analysis section and assisted in revising the manuscript. All authors reviewed and approved the final manuscript, ensuring that it accurately reflects the study’s findings and conclusions. Additionally, all authors agree to be accountable for all aspects of the work, including any questions related to the accuracy or integrity of any part of the work.

## Acknowledgements

The authors would like to thank the study participants and research assistants who collected the data. We would also like to thank EGPAF and the Ministry of Health through the Department of HIV and AIDS for operational guidance on the scope and content of the project to strengthen HIV care and treatment service delivery for people with advanced HIV disease (AHD) and review of this manuscript.

This work was supported by the Bill and Melinda Gates Foundation [INV-004547]. Under the grant conditions of the Foundation, a Creative Commons Attribution 4.0 Generic License has already been assigned to the Author Accepted Manuscript version, which might arise from this submission.

